# Parechovirus infection in early childhood and association with subsequent celiac disease

**DOI:** 10.1101/2020.04.28.20082024

**Authors:** German Tapia, Kateřina Chudá, Christian R. Kahrs, Lars C. Stene, Lenka Kramna, Karl Mårild, Trond Rasmussen, Kjersti S. Rønningen, Ondřej Cinek, Ketil Størdal

## Abstract

**Importance:** Celiac disease is an increasingly common immune-mediated disorder. The potential role of infections in celiac disease development is not well characterized.

**Objective:** To test whether two frequent enteric viruses, parechovirus and anellovirus, were associated with subsequent celiac disease. Our *a priori* hypothesis was that children who later developed celiac disease would have a higher frequency of parechovirus infections before transglutaminase 2 antibody development. Anellovirus testing was exploratory, as a potential marker of immune status.

**Design:** Matched case-control design nested within a longitudinal birth cohort (the MIDIA study) of children at genetic risk for celiac disease.

**Setting:** Children carrying the HLA genotype DR4-DQ8/DR3-DQ2, recruited at birth from the general population throughout Norway during 2001–2007.

**Participants:** Of 220 genetically at-risk children tested for celiac disease, 25 fulfilled the case criteria. Each case was matched for follow-up time, birthdate, and county of residence with two randomly selected children free from celiac disease (controls) from the cohort.

**Exposures:** Parechoviruses, the primary exposure, are infectious agents capable of replication at high virus loads. Anellovirus, previously proposed to reflect immune status, represent a ubiquitous viral exposure at low loads. Viruses were detected and quantified in monthly stool samples (collected from 3 through 35 months of age) using real-time PCR methods.

**Main outcome and measures:** Celiac disease diagnosis according to ESPGHAN 2012 criteria. We retrospectively tested blood samples taken at age 3, 6, 9, and 12 months, and then annually to determine when transglutaminase 2 antibodies developed.

**Results:** Parechovirus was detected in 222 of 2005 stool samples (11.1%), and was more frequent in samples from cases before developing transglutaminase 2 antibodies (adjusted odds ratio [aOR] 1.67, 95% CI 1.14–2.45, P=0.01). The odds ratio was higher when both parechovirus and enterovirus were positive in the same sample (aOR 4.73, 95% CI 1.26–17.67, P=0.02). Anellovirus was detected in 1540 of 1829 samples (84.2%). Anellovirus status did not differ significantly between case and control subjects.

**Conclusions and Relevance:** Parechovirus infections in early life were associated with development of celiac disease in genetically at-risk children, suggesting a novel preventive target if confirmed in future studies.

**Key points:** *Question:* Are parechovirus infections associated with development of celiac disease in childhood?

*Findings:* In this case-control study, nested in a cohort of children genetically at risk for celiac disease, a higher frequency of parechovirus gut infections (tested in monthly stool samples) were associated with later celiac disease. Coinfection with both parechovirus and enterovirus was associated with a markedly increased risk for later celiac disease.

*Meaning:* The association observed between parechovirus and future celiac disease, suggests that these common enteric infections could play a role in celiac disease development.

## INTRODUCTION

Celiac disease (CD) is an immune-mediated disease with known genetic background, known driver (gluten), but unknown environmental triggers.[1, 2] Identifying environmental triggers is challenging, as a subclinical phase detectable by transglutaminase 2 (TG2) antibodies usually precedes clinical CD diagnosis.[3, 4] The subclinical phase of CD can lead to reverse causality (e.g untreated CD might lead to increased infection), unless handled by study design.

One way to minimize reverse causality is using a longitudinal birth cohort, such as our Norwegian MIDIA (Norwegian acronym for *environmental causes of type 1 diabetes*) cohort. MIDIA recruited by HLA genotype predisposing to CD, and was primarily designed to study type 1 diabetes (T1D) pathogenesis. CD and T1D have partly overlapping genetic risk, and could share environmental factors.[5, 6] Infections have long been a proposed environmental factor for T1D,[7] and previous studies suggest that infections may play a role in CD development.[8, 9, 10, 11, 12, 13] We previously performed a metagenomic virome survey of 291 stool samples from MIDIA (manuscript in preparation) to identify candidate human viruses in our population. Assessment of the literature and biological plausibility of the most frequent enteric viruses led us to focus on four major candidate viruses – enteroviruses, adenoviruses, parechoviruses and anelloviruses – which we wanted to study in detail using specific PCR methods. Enterovirus and adenovirus were investigated in a previous study, where we reported an association between human enterovirus and later CD risk.[14] The TEDDY study, a multinational birth cohort including children carrying high T1D risk HLADR-DQ genotypes with a similar longitudinal sampling schedule as MIDIA, recently corroborated the association between enterovirus and CD using a metagenomics approach. [15]

Parechoviruses are enteric viruses closely related to enteroviruses within the *Picornaviridae* family that frequently cause asymptomatic infections in young children.[16, 17] Clinical outcomes of parechovirus infection are uncommon, but can range from mild respiratory disease or gastroenteritis to meningitis, flaccid paralysis or sepsis.[17, 18, 19] An unrelated virus group present in low quantity in a large proportion of samples, the anelloviruses, are persistent human viruses with an unclear relation to disease,[20] but suggested to be associated with autoimmune disorders.[20, 21] Anellovirus is widely disseminated as replication seems linked to the immune system,[20, 22] and could be a marker of immunocompetence.[23, 24, 25]

In this study, we used the MIDIA cohort to investigate if parechovirus and anellovirus frequency differed prior to TG2 antibodies in children later diagnosed with CD (cases) compared with matched controls. We also analyzed if specific genotypes, symptomatic infections or high viral loads were associated with CD, and the influence of enterovirus, infant feeding and symptoms on possible associations.

## METHODS

We designed a nested case-control study of CD (details given in[14]) within a birth cohort originally designed to study T1D (MIDIA, described in[26]). Figure 1 outlines the study design and formation of the study sample. Characteristics of participants (shown in Table 1) were obtained from MIDIA questionnaires as described by Stene et al.[26] unless noted otherwise. MIDIA and the CD sub-study were approved by The Regional Committee for Medical Research ethics.

**Figure 1:**
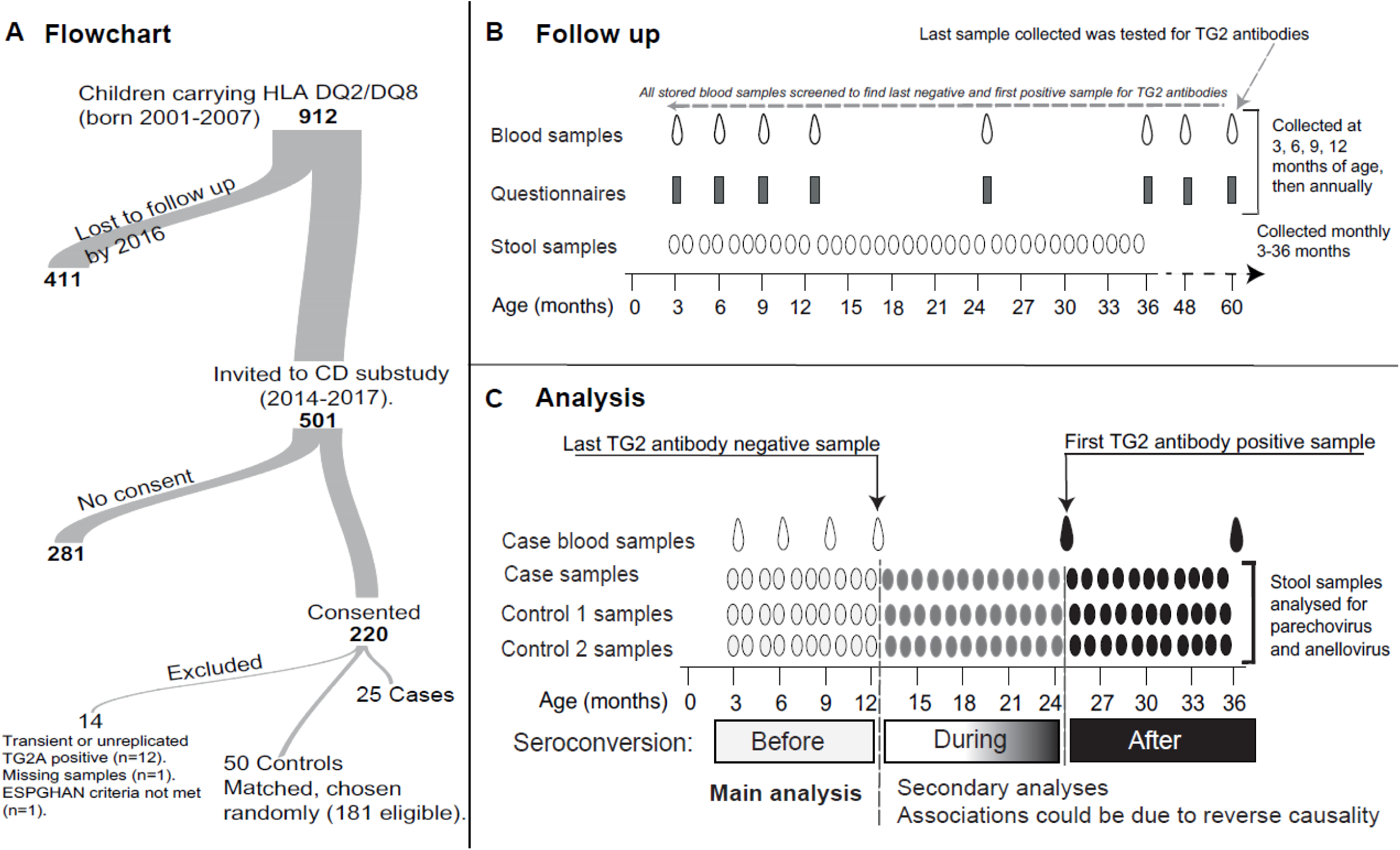
Study design. A flowchart of the current study (A). An illustration of the follow-up and TG2 antibody testing in the study (B). An illustration of the main and secondary analyses in the study (C). R (version 3.6.1, R core team 2018), package riverplot[43] was used in the generation of panel A.

**Table 1.** Characteristics of the study participants

|  | **Cases (n=25)** | **Controls (n=49)** |
| --- | --- | --- |
| Girls (n (%)) | 16 (64) | 26 (53) |
| Other children in household (n (%))<sup>0</sup> |  |  |
| *None* | 7 (28) | 15 (31) |
| *1* | 10 (40) | 28 (57) |
| *≥2* | 8 (32) | 6 (12) |
| Islet autoimmunity (n (%))<sup>a</sup> | 4 (16) | 1 (2) |
| Family history of celiac disease<sup>b</sup> | 7 (28) | 4 (8) |
| Mean (SD) age (months) at antibody screening test<sup>c</sup> | 99 (30) | 106 (21) |
| Mean (range) age (months) of last antibody negative sample | 30 (6-75) | n.a |
| Mean (SD) age (months) at end of study<sup>d</sup> | 119 (19) | 119 (19) |
| Mean (SD) age (months) at last stool sample | 32 (7) | 34 (3) |
| Stool samples<sup>e</sup> | 703 | 1458 |
| Parechovirus |  |  |
| *Stool samples with parechovirus data (n (%))* | 647 (92) | 1358 (93) |
| *Parechovirus positive<sup>f</sup> stool samples (n (%))* | 84 (13.0) | 138 (10.2) |
| *Median (range) count of positive samples per child* | 3 (1 – 6) | 3 (0 – 8) |
| Anellovirus |  |  |
| *Stool samples with anellovirus data (n (%))* | 588 (85) | 1241 (84) |
| *Anellovirus positive<sup>f</sup> samples (n (%))* | 506 (86) | 1034 (83) |
| *Median(range) count of positive samples per child* | 22 (5 – 31) | 21 (10 – 32) |

<sup>0</sup> No children in the matched case-control set were cousins, half siblings or siblings to each other.
<sup>a</sup> Determined as described in Stene et al.[26]
<sup>b</sup> Known celiac disease in first degree relative or half-sibling ascertained at celiac disease screening 2014-2016.
<sup>c</sup> Antibody screening test performed on the most recent blood sample from both case and control children at time of inclusion in celiac disease sub-study of MIDIA
<sup>d</sup> At time when all 25 cases were ascertained by end of February 2016.
<sup>e</sup> Discrepancy between total number of stool samples, and samples providing virus data was due to missing samples or failed laboratory test.
<sup>f</sup> Positivity defined as $\geq 10$ copies of viral genomes per $\mu\text{l}$

Briefly, during 2001–2007, 46 939 newborns throughout Norway were screened for the HLA-DQ2/DQ8 genotype (HLA-DRB 1*04:01-DQA1*03-DQB 1*03:02/DQA1*05-DQB 1*02) conferring an increased risk of T1D as well as CD. This genotype was identified in 912 (1.9%) children, who were followed with repeated questionnaires, plasma samples (collected at ages 3, 6, 9, 12 months, then annually), and stool samples (monthly, ages 3–35 months). Children actively participating during 2014–2016 (n=501) were invited to CD screening (n=220 consenting, Figure 1A). Those consenting tended to have higher prevalence of islet autoimmunity, fewer siblings, and slightly higher prevalence of CD family history at recruitment. Children with known CD (n=13) were identified by parental questionnaire, followed by review of medical records. Consenting participants were screened for TG2 antibodies (using EliA Celikey IgA/EliA GliadinDP IgG, Thermo Fischer Scientific, Phadia AB; Uppsala, Sweden) in their most recent blood sample, detecting 12 additional children after diagnostic workup (in total 25 children with CD). The CD diagnosis was based on ESPGHAN 2012 criteria. Briefly, these criteria require a biopsy with Marsh grade 2-3, or TG2-titers >10x the normal upper limit, presence of endomysial antibodies in a separate blood sample and a HLA risk genotype. [27] Two controls per case were matched for follow-up duration, date of birth, and county of residence. One matched control was excluded owing to missing stool samples. Stored plasma samples from cases were screened for TG2 antibodies and antibodies against deamidated gliadin peptides (DGP), as per guidelines at the time, in order to identify the last negative and first positive sample for CD antibodies. The same testing was performed in controls to confirm the absence of CD. Children who were transiently positive for TG2 antibodies did not fulfill the case definition and were excluded as controls, with new random controls picked if necessary. No children were diagnosed based on DGP, and in practice TG2 antibodies was sufficient alone, with TG2 antibodies also defining the seroconversion interval. In the 25 case-control groups, 15 children with CD seroconverted before the end of stool sampling (35 months of age), while the remaining seroconverted at a later age.

### Detection of parechovirus and anellovirus in stool samples

The studied viruses were *Human parechovirus* (species *Parechovirus* A; genus *Parechovirus*; family *Picornaviridae*, referred to as parechovirus hereinafter, a single strand positive (ss+)RNA virus), and the genera *Alpha- and Beta-* and *Gammatorquevirus* of the family *Anelloviridae* (referred to as anellovirus hereinafter, circular, negative sense, ssDNA viruses). As anellovirus is characteristic by its extreme genetic diversity, it was not investigated at species level or sequenced.[28] All available fecal samples from cases and controls were subjected to RNA and DNA extraction using Qiagen chemistry (Qiagen, Hilden, Germany). Parechovirus was tested (n=2005 samples) with real-time RT-PCR using primers based on Corless et al.[29] and a probe redesigned to detect novel parechovirus types. Anellovirus was tested (n=1829 samples) using real-time PCR adopted from Thom et al.[30]. Differences in number of tested samples was due to sample exhaustion. The positivity threshold was set to 10 copies/μL. The VP3-VP1 gene junction of parechovirus samples with ≥10 copies/μL was genotyped with next generation amplicon[31] or Sanger[32] sequencing. Oligonucleotide sequences used and details are given in the online-only supplement and eTable 1. Detailed protocols are available upon request.

### Statistical analysis

#### Main analysis

We analyzed the association of virus with CD primarily by using a mixed effects logistic regression model with random intercepts for each child and for each matched set to account for the matched design. Virus positivity was the dependent variable (each virus a separate model) and case-control status an independent variable. The odds ratio (OR) for CD is then interpreted as the odds of a sample being virus positive given that it came from a case, compared with the same odds for virus positivity for samples from matched controls. Potential predictors of viral infection and CD were included as covariates in the main regression model: sex, age, age squared (to accommodate established non-linear associations between age and virus positivity), sample month, siblings (categorized as none, 1, or ≥2), and CD family history. The primary analysis included only samples collected before the last sample negative for TG2 antibodies (Figure 1C), to avoid potential reverse causality. The analyses were run in Stata, release 15 (StataCorp, College Station, Texas).

#### Pre-planned secondary analyses

We additionally adjusted the primary analysis for enterovirus positivity, islet autoimmunity, age at gluten introduction and breastfeeding duration, and stratified the analysis using only samples collected while breastfeeding or after weaning, and only before, after or during (±1 month) gluten introduction. We analyzed the association between CD and parechovirus types present in ≥20 samples. We investigated the association between virus and CD for stool samples collected during, and after development of TG2 antibodies. During seroconversion, we cannot determine if infections were prior or after appearance of TG2 antibodies. Analyzes after seroconversion was to determine if development of TG2 antibodies influenced infection frequency. We analyzed if high virus quantity (≥median quantity of positive samples), longer viral shedding (≥2 consecutive monthly samples), or symptomatic infections were associated with later CD. Data from our earlier study reporting an enterovirus-celiac disease association [14] was used to analyze possible confounding or mediation by enterovirus of the observed parechovirus-CD association and coinfections (samples positive for both viruses set as positive; other non-missing samples were set as negative, otherwise as the main analysis). To statistically adjust parechovirus results with our earlier data on enterovirus, we ran a conditional logistic regression including only data prior to development of TG2 antibodies with CD as outcome and number of parechovirus positive as exposure, adjusting for number of enterovirus positive samples, matching variables and covariates in the main analysis.

#### Exploratory analyses

As enterovirus and parechovirus are both picornaviruses, we combined these into one variable (positive for enterovirus or parechovirus), which was analysed as the main analysis. Recently, the TEDDY study reported results from a metagenomics survey, where infections were analysed by year of life (1^st^ and 2^nd^). [15] To attempt to replicate these results, we investigated the association between infections (enterovirus, parechovirus, adenovirus, and anellovirus) the first or second year of life and later celiac disease. These analyses was restricted to samples collected after the introduction of dietary gluten, and time of TG2 antibody development was set to the first positive sample (instead of the last negative, as otherwise in our manuscript) to replicate the TEDDY approach.[15] These analyses were adjusted for calendar month of sampling using sine and cosine terms, [33] due to low number of samples in these sub-analyses, but otherwise adjusted as the main analysis.

## RESULTS

### Parechovirus is a frequent childhood viral exposure

Parechovirus was detected in 222 of 2005 (11.1%) tested stool samples in total, and was positive at least once in 72 out of 74 (97.3%) tested children. The highest parechovirus quantity was 2.5×10^6^ copies/μl (median 1,979 copies/μl in samples with ≥10 copies/μl). Parechovirus was more common during September–January (eFigure 1), first encountered in early life (eFigure 2), and most frequent at 9–18 months of age (eFigure 3). The most common type was parechovirus 1 (n=133, 62.1%), followed by 6 (n=55, 30.2%), 3 (n=21, 11.5%) and 4 (n=1, 0.6%). A phylogenetic tree of parechovirus sequences is shown in eFigure 4. The percentage of samples available at each collection age for this study are shown in eFigure 5.

### Parechovirus was more frequent in case children prior to TG2 antibodies

Before development of TG2 antibodies the frequency of parechovirus positive samples was higher among cases (63/392 samples, 16.1%) than for matched control children (87/787 samples, 11.1%), adjusted OR (aOR) of 1.67, 95% CI 1.14–2.45, P=0.01 (Table 2). Parechovirus 1 and 6 were more frequent in cases (Table 2). During or after development of TG2 antibodies, the frequency of parechovirus RNA in stool samples did not differ by case status (Table 2). Parechovirus positive samples concurrently (±15 days) of parent-reported diarrhea were more frequent in cases (6/384, compared to 5/780 samples in controls) prior to TG2 antibody development (aOR 3.90, 95% CI 1.06–14.29, P=0.04), but parechovirus positive samples with reported fever or common cold were not (data not shown). The frequency of high quantity or long-lasting parechovirus infections did not differ between case and control children (Table 2).

**Table 2.** Association between parechovirus and celiac disease^a^

| Parechovirus | Cases<br>(n=25) | Control<br>s<br>(n=49) | Unadjusted | Adjusted<sup>b</sup> |  |
| --- | --- | --- | --- | --- | --- |
|  | *Positives (n)/Total*<br>*(N) samples (%)* |  | **Odds ratio**<br>**(95% CI)** | **Odds ratio**<br>**(95% CI)** | **P**<br>**value** |
| **Main analysis<sup>c</sup>** |  |  |  |  |  |
| Parechovirus, before TG2 antibody seroconversion | 63/392<br>(16.1%) | 87/787<br>(11.1%) | 1.57 (1.07 - 2.28) | 1.67 (1.14 - 2.45) | 0.01 |
| **Secondary analyses** |  |  |  |  |  |
| Parechovirus, during TG2 antibody seroconversion | 12/136<br>(8.8%) | 31/320<br>(9.7%) | 0.89 (0.44 - 1.81) | 1.12 (0.48 - 2.60) | 0.79 |
| Parechovirus, after TG2 antibody seroconversion | 9/119<br>(7.6%) | 20/251<br>(8.0%) | 0.96 (0.42 - 2.22) | 1.04 (0.40 - 2.70) | 0.94 |
| Parechovirus 1, before TG2 antibody seroconversion | 31/360<br>(8.6%) | 47/747<br>(6.3%) | 1.40 (0.88 - 2.25) | 1.42 (0.84 - 2.39) | 0.19 |
| Parechovirus 6, before TG2 antibody seroconversion | 18/347<br>(5.2%) | 22/722<br>(3.0%) | 1.80 (0.87 - 3.73) | 1.87 (0.88 - 3.97) | 0.10 |
| Parechovirus, long<sup>d</sup> infections before TG2 antibody seroconversion | 21/350<br>(6.0%) | 30/730<br>(4.1%) | 1.49 (0.84 - 2.64) | 1.54 (0.83 - 2.86) | 0.18 |
| Parechovirus, high<sup>e</sup> quantity before TG2 antibody seroconversion | 33/392<br>(8.4%) | 48/787<br>(6.1%) | 1.42 (0.89 - 2.24) | 1.41 (0.85 - 2.35) | 0.19 |

<sup>a</sup>The number of positive/total stool samples included in each analysis (for the cases and controls) and the results from the mixed effects logistic regression. In total, there were 222 positive samples, with 150 before, 43 during and 29 after TG2 antibody seroconversion. In the analysis for the specific genotypes, samples positive for other parechovirus types have been set to missing.
<sup>b</sup>Adjusted for sex, age, age squared, month of sample collection, number of siblings, and family history of celiac disease.
<sup>c</sup>Before: prior to last TG2 antibody negative blood sample. During: between the last TG2 antibody negative and the first TG2 antibody positive blood sample. After: after the first TG2 antibody positive blood sample.
<sup>d</sup>Defined as infections lasting $\geq 2$ months
<sup>e</sup>Defined as ≥ median quantity ( $\geq 1979$ copies/ μl) of parechovirus positive samples

### Parechovirus and enterovirus were independently associated with later CD

Coinfection with enterovirus and parechovirus was observed in 10/427 case vs. 5/845 control samples (aOR 4.73, 95% CI 1.26–17.67, P=0.02). Adjusting for enterovirus in our main analysis gave similar estimates (aOR 1.81, 95% CI 1.23–2.67, P=0.003), and adjusting our earlier reported enterovirus-celiac disease association for parechovirus did not substantially change our published results (aOR 1.76, 95%CI 1.24–2.50, P=0.001). Using a conditional logistic regression model, parechovirus estimates did not change after statistical adjustment for enterovirus (aOR 1.71, 95% CI 1.04–2.80 vs. aOR 1.69, 95% CI 1.03–2.78 when additionally adjusted for enterovirus), suggesting independent associations. We assessed potential deviation from linearity by categorizing into approximate tertiles (0, 1–2, and ≥3 parechovirus positive samples before TG2 antibodies), in the conditional logistic regression model and found no evidence for deviation of additivity in the log odds (data not shown). Combining enterovirus and parechovirus into one variable (positive for either) gave similar results as in the main analysis (aOR 1.61, 95%CI 1.19 – 2.17, p = 0.002).

### Secondary analyses did not substantially influence interpretation of results

Adjusting our main analysis for breastfeeding, age at gluten introduction or islet autoimmunity did not change our results substantially (data not shown). Parechovirus infections after gluten introduction (median age 6 months, range 2–10), and while breastfed (median duration 12 months, range 1–23) were more frequent in case children (eTable 2).

### Anellovirus was asymptomatically present in a large majority of stools

Anellovirus was present in all children, being positive in 1540 of 1829 (84.2%) fecal samples. The longest continuous shedding of anellovirus was 32 months, and maximum quantity was 238,773 copies/μl (median 543 copies/μl in positive samples). Anellovirus positivity did not show seasonality (data not shown) and was highly prevalent at all ages (eFigure 3).

### Anellovirus frequency was not associated with seroconversion for TG2 antibodies

The frequency of anellovirus positive stool samples before or after development of TG2 antibodies did not differ significantly by case status. During development of TG2 antibodies (between the last negative and first positive TG2 antibody measurement), there was a borderline association, with cases having a higher frequency of positive samples (92.5% vs 83.4%, aOR 3.32, 95% CI 1.01–10.92, P=0.05, Table 3). The frequency of anellovirus samples with high quantity did not differ between cases and controls (Table 3). Long-lasting anellovirus infections were not separately analyzed due to their persistent nature.

**Table 3.** Association between anellovirus with celiac disease^a^

| **Anellovirus** | **Cases**<br>**(n=25)** | **Control**<br>**s**<br>**(n=49)** | **Unadjusted** | **Adjusted<sup>b</sup>** |  |
| --- | --- | --- | --- | --- | --- |
|  | *Positives (n)/Total*<br>*(N) samples (%)* |  | **Odds ratio**<br>**(95% CI)** | **Odds ratio**<br>**(95% CI)** | **P**<br>**value** |
| **Main analysis<sup>c</sup>** |  |  |  |  |  |
| Anellovirus | 331/376<br>(88.0%) | 645/758<br>(85.1%) | 1.28 (0.61 - 2.71) | 1.39 (0.64 - 3.03) | 0.40 |
| **Secondary analyses** |  |  |  |  |  |
| Anellovirus, high<sup>d</sup> quantity before TG2 antibodies | 188/376<br>(50%) | 325/758<br>(42.9%) | 1.37 (0.51 – 0.94) | 1.41 (0.92 – 2.15) | 0.11 |
| Anellovirus, during TG2 antibody seroconversion | 99/107<br>(92.5%) | 216/259<br>(83.4%) | 2.50 (0.79 - 7.86) | 3.32 (1.01 - 10.92) | 0.05 |
| Anellovirus, after TG2 antibody seroconversion | 76/105<br>(72.4%) | 173/224<br>(77.2%) | 0.75 (0.32 - 1.78) | 0.92 (0.36 - 2.36) | 0.86 |

<sup>a</sup> The number of positive/total stool samples included in each analysis (for the cases and controls) and the results from the mixed effects logistic regression are shown. In total, there were 976 positive samples prior to, 315 during and 249 after TG2 antibody seroconversion.
<sup>b</sup> Adjusted for sex, age, age squared, month of sample collection, number of siblings, and family history of celiac disease.
<sup>c</sup> Before: prior to last celiac disease antibody negative blood sample. During: between the last celiac disease antibody negative and the first celiac disease antibody positive blood sample. After: after the first celiac disease antibody positive blood sample.
<sup>d</sup> Defined as ≥ median quantity (543 copies/ μl) in anellovirus positive samples

Anellovirus infections prior to, at the same time, or after gluten introduction, or while breastfed or after weaning, did not differ according to case status (eTable 2). The frequency of anellovirus positive samples with concurrent symptoms did not differ between case and control children (data not shown).

### Exploratory analyses stratified by year of life

Samples positive for parechovirus were more common in case children the first year of life (aOR 3.09, 95% CI 1.02 – 9.39, p = 0.05), and samples positive for enterovirus were more common in case children the second year of life (aOR 2.08, 95% CI 1.33 – 3.27, p = 0.001). Anellovirus and adenovirus did not show any statistically significant association with case status in the first or second year of life (eTable 3).

## DISCUSSION

Case children were more likely to have parechovirus positive samples prior to the appearance of TG2 antibodies. This association did not seem to be confounded by enterovirus, or other variables considered.

### Strengths and limitations of the study

The main strengths of the study are the longitudinal nature of the study and the careful construction of the case-control dataset. Screening of TG2 antibodies coupled with medical review and diagnostic workup, with widely accepted CD diagnostic criteria used for case definition makes the possibility of misclassification small. Testing for TG2 antibodies in stored blood samples allowed us to determine seroconversion interval and investigate infections prior to TG2 antibody development, thus minimizing potential spurious associations due to reverse causation. We observed no association with parechovirus positivity after TG2 antibody development, implying that CD autoimmunity does not influence parechovirus frequency. Another strength is how candidate viruses were selected. Enteric human candidate viruses were identified for quantitative PCR testing using metagenomic virome sequencing, ensuring that we could focus on frequent viruses present in the time period studied. We excluded less frequent viruses which could result in insufficient statistical power, and viruses without a plausible link to CD which could lead to multiple testing problems. PCR was the method of choice for assessing the viral exposure, as metagenomic sequencing is presently unsuitable for generating truly quantitative data. We adjusted for potential confounders, and are not aware of strong confounders neglected.

There are several limitations to this study. We lack data on host immune responses, such as cytokine and virus-specific antibody levels, which could be relevant for disease pathogenesis. Although we found statistically significant associations, and included a large number of samples, the modest number of children with CD led to limited precision in estimates. The smaller sample size in sub-analyzes must be kept in mind when evaluating these. Children in the MIDIA study all carry the HLA DR3-DQ2/DR4-DQ8 genotype, and we cannot generalize our results to subjects with other genotypes. However, DR3-DQ2 and DR4-DQ8 are the main CD HLA susceptibility genotypes, with the majority of CD cases carrying at least one of these. Children were followed up for ~10 years, and some could seroconvert or be diagnosed later, but most children are expected to develop CD antibodies by ten years of age.[34] All children were screened, and no control child developed CD antibodies during the study. As any observational study, we cannot rule out unmeasured or residual confounding, necessitating further prospective studies to replicate findings.

### Comparison with other studies

One small prior study investigating parechovirus and CD did not detect any parechovirus measured in stool at 3 and 6 months of age.[35] In our data, parechovirus prevalence was very low in this age-group (e.g. 1.0% at ≤4 months of age) which would limit statistical power for analyses restricted to this age-group.

We have previously reported an association between enterovirus and later CD.[14] A similar association with enterovirus was also recently reported in the second year of life in a large metagenomic study, which did not include parechovirus data.[15] To attempt to replicate results from the TEDDY study,[15] we did an exploratory analysis stratifying our analysis into the first and second year of life. We could not replicate the association reported between adenovirus infections the first year of life and later celiac disease autoimmunity.[15] Parechovirus was more frequent in samples from case children the first year of life, and enterovirus was more frequent in samples from case children the second year of life. This could imply there are different risk periods for different viruses, or differences could be due to diet or development. Parechovirus and enterovirus are two of the most common human picornavirus genera, with overlapping presentation and seasonality, and both primarily replicate in epithelial cells of the intestinal mucosa and in adjacent lymphoid tissues.[17] As these viruses could presumably share infection risk factors, we tested whether our results were influenced by enterovirus, but parechovirus remained significantly associated with CD after adjustment for enterovirus. Interestingly, coinfection with both viruses associated with a highly increased CD risk, and both enterovirus and parechovirus presenting with diarrhea had increased risk estimates.[14]

There are no previous studies investigating anelloviruses and CD, but the ubiquitous nature of anelloviruses makes the disease association difficult to interpret.[20] Anelloviruses are believed to replicate in stimulated peripheral blood mononuclear cells, so the presence or level of anellovirus could be a marker of already present disease or infection.[20, 22, 36, 37, 38, 39] A large majority of samples were positive for anellovirus. Yet, we cannot rule out that anellovirus could be a marker of underlying pathology, or disease progression, as in this study their occurrence was marginally increased during TG2 antibody seroconversion.

Studies on specific viral species have implicated members of *Reoviridae*[10, 13] and *Picornaviridae*[14, 15] in CD etiology. These observations and the present results could be interpreted to mean that infections with RNA viruses are associated with CD development, either in general, in these viral families, or in specific viral species/types/strains. Risk estimates varied between parechovirus types in our study, which could imply some - yet statistically insignificant - differences between parechovirus types regarding CD etiology, similar to the one shown for reovirus types in mouse models.[13] Picornavirus infections have been hypothesized to be involved in autoimmunity,[40] and could contribute to CD development in several ways. Infections could disrupt the mucosal barrier, leading to translocation of gluten peptides into the mucosa,[41] tissue injury and/or inflammation which could provide signals necessary for breaking tolerance against gluten peptides, and/or activate gluten-specific CD4^+^ T-cells.[2] Infection with RNA viruses could activate extracellular TG2,[42] leading to increased gluten deamidation at the site of a viral infection, where it could be taken up, processed and presented by antigen-presenting cells concurrently with viral proteins. Viral infections are thus biologically plausible environmental factors, but mechanistic studies are needed to elucidate potential mechanisms.

### Implications, Conclusions and future work

These results could imply that parechovirus infections are associated with increased risk of CD. Picornavirus infections could be one of several environmental factors associated with increased risk for development of CD. Our findings suggest novel preventive targets if corroborated in future studies.

## Data Availability

Sequences from this study are deposited in GenBank, accession numbers MT009645-MT009897.

## ACKNOWLEDGEMENTS

**Author contributions:** Conceived and designed the study: CRK GT OC LCS KS. Virus detection and genotyping: OC KC LK. Analyzed the data: GT LCS. Contributed reagents/materials/analysis tools: GT OC KS. Drafted the first version of the paper: GT. Reviewed, commented and revised the paper: All authors. **Data access:** Sequences from this study are deposited in GenBank, accession numbers MT009645-MT009897. **Disclosures:** The authors have no conflicts of interest relevant to this article to disclose. **Grant support:** This work was supported by The Research Council of Norway (grant numbers 205086/F20 to GT, 166515/V50 to KSR), and the Project for the Conceptual Development of Research Organization 00064203 (University Hospital Motol, Prague, Czech Republic). Kateřina Chudá has been supported with funding from the European Union Horizon 2020 research and innovation programme under grant agreement No 874864 HEDIMED. Christian Riddervold Kahrs has been supported with grants from Østfold Hospital Trust, Kalnes, Norway. The EliA Celikey and EliA Gliadin kits were supported by Thermo Fisher Scientific, Norway. The funders had no role in study design, data collection and analysis, decision to publish, or preparation of the manuscript. **Acknowledgments:** We express our sincere gratitude to the participants in the MIDIA cohort. We also especially thank Nicolai Andre Lund-Blix of Oslo University Hospital, Norway, for his help with the breastfeeding data and discussions; Lisa Merete Markussen of Akershus University Hospital for help with analyzing celiac disease antibodies in historical blood samples; Kaja Klykken Aas and Nina Stensrud from the Norwegian Institute of Public Health for handling plasma and stool samples in the MIDIA project; and the public health nurses in the MIDIA project (Asbjørg Skorge Hornseth, Liv Kjeldstadli Onsrud, and Turid Wetlesen).

